# Deep Learning-Based Universal Expert-Level Recognizing Pathological Images of Hepatocellular Carcinoma and beyond

**DOI:** 10.1101/2020.03.22.20041178

**Authors:** Wei-Ming Chen, Min Fu, Cheng-Ju Zhang, Qing-Qing Xing, Fei Zhou, Meng-Jie Lin, Xuan Dong, Qi-Zhong Zheng, Mei-Zhu Hong, Jin-Shui Pan

## Abstract

**Purpose:** Human-based medical-image interpretation always falls into the predicament between specialized practitioners and expanding medical imaging. We aim at developing a diagnostic tool for pathological-image classification by using transfer learning that can be applied to diverse tumor types.

**Experimental Design:** In this study, images were retrospectively collected and prospectively analyzed using machine learning. Microscopic images of liver tissue that show or do not show hepatocellular carcinoma were used to train and validate a classification framework based on convolutional neural network. To evaluate the universal classification performance of the artificial-intelligence (AI) framework, histological images from colorectal tissue and breast were also collected. Training and validation set of images were collected from Xiamen Hospital of Traditional Chinese Medicine whereas test set of images were collected from Zhongshan Hospital Xiamen University.

**Results:** Accuracy, sensitivity, and specificity were reported and compared to human image interpretation and other AI image classification systems such as AlexNet and GoogLeNet. For the test dataset, sensitivity, specificity, and area under the curve of the AI framework were 99.1%, 98.0%, and 0.960, respectively. In human-machine comparisons, the accuracy of the AI framework was 98.5%, while the accuracy of human experts fluctuated between 93.0% and 95.0%. Based on transfer learning, the AI framework accuracy for colorectal carcinoma, breast invasive ductal carcinoma, were 96.8%, and 96.0%, respectively.

**Conclusions:** The performance of the proposed AI framework in classifying histological images with hepatocellular carcinoma is comparable to the classification by human experts. With limited training, the proposed AI framework has potential universality in histological image classification.

**Study Highlights:** WHAT IS KNOWN

✓ Accurate recognition of medical images is the basis for clinical decision-making.
✓ Unresolved challenge exists between specialized practitioners and expanding medical imaging output.

WHAT IS NEW HERE

✓ Proposed AI framework has excellent performance in classifying hepatocellular carcinoma.
✓ The AI framework has universal feature in classifying other types of histological images.
✓ The AI framework may help to interpret emerging imaging technology.

## Introduction

Hepatocellular carcinoma (HCC) ranks as the fifth most common cancer worldwide and the second most common cause of cancer-related deaths.(1) In the United States and China, HCC is estimated to be the fourth and third most common cause of cancer-related deaths, respectively.(2, 3) This type of liver cancer usually develops in patients with liver cirrhosis, especially in chronic hepatitis B (CHB) or chronic hepatitis C (CHC)-related liver cirrhosis.(4-6) In cirrhosis cases, numerous nodules of varying sizes are found in the liver. Benign and malignant intrahepatic nodule identification is often a huge challenge for computed tomography (CT) or magnetic resonance imaging (MRI)-based diagnosis. Definitive diagnosis of intrahepatic nodules usually depends on liver biopsy. Histopathology diagnosis is the gold standard for determining the nature of hepatic space occupying lesions. However, diagnosing a large number of pathology slide images is laborious. Additionally, substantial observer-to-observer variation in liver biopsy assessments cannot be neglected.(7) Another challenge faced during medical-image diagnostics is the patient-to-patient variability in the pathology of disease manifestation. Even experienced pathologists differ significantly in the interpretation of histopathology of the same disease. Therefore, it is necessary to develop novel auxiliary diagnostic facilities.

Diagnostic approaches to HCC include ultrasound, CT, and MRI, with histopathology diagnosis as the “gold standard”.(8) In addition to HCC, the colorectal cancer (CRC) and breast invasive ductal carcinoma (BIDC), described later, are some of the most common tumors. In 2015, CRC was estimated to be the fifth most common cause of cancer-related deaths in China.(3) Similarly, according to the 2018 United States cancer statistics published by Siegel et al.,(2) CRC ranks as the third most common cause of cancer-related deaths in both men and women. Moreover, adenocarcinoma is the most common type of CRC, making a confirmed diagnosis of CRC dependent on pathology imaging and interpretation. Diagnostic methods of CRC include CT, colonoscopy and subsequent tissue examination,(9) while breast cancer diagnostics include ultrasound and mammography.(10) In 2015, breast cancer was estimated to have contributed toward most new cancer cases in China.(3) The same was true in the United States, according to cancer statistics published by Siegel et al.(2) Invasive ductal carcinoma is the most common type of breast cancer that is diagnosed histologically.(11) Similar to CRC, histopathology examination is also the diagnostic gold standard for BIDC. To summarize, there are numerous imaging diagnostic means for almost all diseases. More than that, almost all diagnostic imaging can produce a large number of medical images. For example, a plain scan combined with contrast-enhanced CT or MRI can produce more than 1000 images per examination, while capsule endoscopy can produce more than 40000 medical images per examination. Thus, interpretation of these images can be time-consuming.

With the continuous emergence of new technologies, the number and types of medical images have expanded at an unprecedented rate. However, an unresolved challenge exists due to the imbalance between the ability and number of specialized practitioners and expanding medical imaging output. Usually, a physician is familiar with only a few or even just one type of diagnostic imaging, whereas the interpretation of countless medical images requires human expertise and judgement to correctly understand and triage. Therefore, it is undoubtedly attractive to develop an artificial intelligence (AI) system of high classification accuracy with universal recognition capability.

In recent years, AI has been widely used in various fields.(12-14) For classification that is difficult for human experts or where rapid review of an immense number of images is needed, AI has outstanding advantages and has a revolutionary role in disease diagnosis. In the present study, we developed an effective convolutional neural network (CNN) based on a deep-learning algorithm to classify medical images. We also evaluated the generalizability performance of the proposed AI system in interpreting histological images of several common types of tumors through transfer learning.

## Methods

### Patients

Patients who underwent biopsy or surgical resection due to diseases of the liver, colorectum, or breast in Xiamen Hospital of Traditional Chinese Medicine, or Zhongshan Hospital Xiamen University between June 1, 2010 and December 31, 2017. Adult patients who aged between 18 and 75 were enrolled. The enrolled participants had biopsy or surgical resection specimens with completed structure, and one of the following conditions: 1) chronic hepatitis-related to hepatis B virus (HBV) or hepatis C virus (HCV), without HCC, with or without liver cirrhosis; 2) HCC companied by HBV-related or HCV-related chronic hepatitis, with or without liver cirrhosis; 3) CRC; 4) BIDC. All of HCC, CRC, and BIDC were further confirmed on the basis of surgically resected specimens. Necroinflammatory activity and fibrosis or cirrhosis related to chronic hepatitis were recorded using the Scheuer system.(15) Histological diagnosis of HCC or CRC was made according to the digestive tumor-classification system formulated by the World Health Organization (WHO) in 2010 (16) while the histological diagnosis of BIDC was made according to the breast-tumor-classification system formulated by WHO in 2012.(17) There were no exclusion criteria regarding gender, or race. This study was approved by the Ethics Committees of Zhongshan Hospital, Xiamen University. Written informed consent was waived by the Ethics Commission of the designated hospital because of non-interventional study and no identifiable personal information was recorded.

### Images

Collected tissue samples were fixed in the wax, followed by slicing at the thickness of 3 μm, and then staining by hematoxylin-eosin. After the process, histological images were collected with a 200-fold magnification. Two to 6 images were collected for each patient, and there was no overlap between the images. For a case with tumor, an image of tumor was collected companied by corresponding non-tumor image, which was taken at 2 cm away from tumor focus. Strategy of “full field” was adopted. An image of tumor was taken in the focus of tumor whereas an image of non-tumor was taken in the focus non-tumor. In other words, the entire image of tumor was consisted of tumor tissue while the whole image of non-tumor was consisted of non-tumor tissue. Each image was examined by a panel of two independent pathologists, each with over 15 years of pathology experience. If there was a disagreement in clinical labeling, the image was further arbitrated by a panel of senior pathology specialists. Before training the AI framework, a whole image was dissected into partial regions of 1920 × 1280 pixels. Identifiable personal information, such as name of the enrolled patients, name of hospital, etc, was removed.

### Datasets

Histological images collected from Department of Pathology, Xiamen Hospital of Traditional Chinese Medicine acted as training and validation sets while histological images collected from Department of Pathology, Zhongshan Hospital Xiamen University acted as test sets to further verify the performance in classifying. Histological images of HCC, non-HCC, CRC, non-CRC, BIDC, and non-BIDC were collected independently from these two hospitals in the same way.

### Training and validation the AI algorithm

Each divided image of 1920 × 1280 pixels was imported into the database with multiple layers of classification. For an entire image, it would be classified as “tumor” if tumor was identified even in only one dissected image. However, only when all of dissected images were recognized as “non-tumor” would the entire image be classified into the group of “non-tumor”. Collected liver pathology images were randomly divided into training set and validation set at a ratio of 3:1. The training set was used to train the AI algorithm whereas the validation set was employed to evaluate the classification performance of the trained AI algorithm. This process was repeated five times.

Based on deep learning, we developed an AI algorithm, and used the PyTorch platform to adopt the ResNet-34 architecture pretrained using the ImageNet dataset.(18) The retraining consisted of initialization of the convolutional layers with loaded pretrained weights and updating of the neural network to recognize our classes such as HCC or non-HCC. In the training process, the network structure was kept unchanged. However, the network learning rates, except the last fully connected layer, were tuned to be 0.001. The learning rate of the last fully connected layer was tuned to be 0.02 (0.001 × 20), and weights were updated using backpropagation. This strategy tended to update the first several layers slowly, while updating the output layer more efficiently. Layer training was performed by stochastic gradient descent in batches of 64 images per step by using stochastic gradient descent optimizer. The training procedure was run for 25 epochs with a dropout ratio equal to 0.5. The modified ResNet-34 was trained on an 14.04.1-Ubuntu computer with Intel(R) i7-5930K CPU @ 3.50 GHz. An NVIDIA GTX 1080Ti 11 GB GPU was utilized to accelerate training.

### Testing of the AI algorithm

After the training process was finished, histological images collected from Department of Pathology, Zhongshan Hospital Xiamen University was used as test set to monitor the classifying decisions of the trained algorithm.

### Comparison between the AI algorithm and human experts

Histological images collected from Department of Pathology, Zhongshan Hospital Xiamen University were also sent to expert pathologist to make a diagnosis.

Classification performance was compared with that of the AI algorithm. Expert pathologists were senior staffs of Department of Pathology, Zhongshan Hospital Xiamen University, with clinical experience about 15 years. Diagnosis was made independently. The error rates were determined for the AI algorithm and for each of the human experts. Comparison was also conducted between the proposed AI algorithm and other framework such as AlexNet and GoogLeNet.(19, 20)

### Transfer Learning of the AI algorithm

Transfer learning was developed by Donahue et al.(21) To evaluate the transfer-learning performance, the trained AI algorithm was further tested by two other kinds of tumors: CRC and BIDC. The classifying performance of the trained AI algorithm was determined independently in each kind of tumor.

The study design was shown in Figure 1 while the CNN schematic for HCC classification was shown in Supplementary Figure 1.

**Figure 1.**
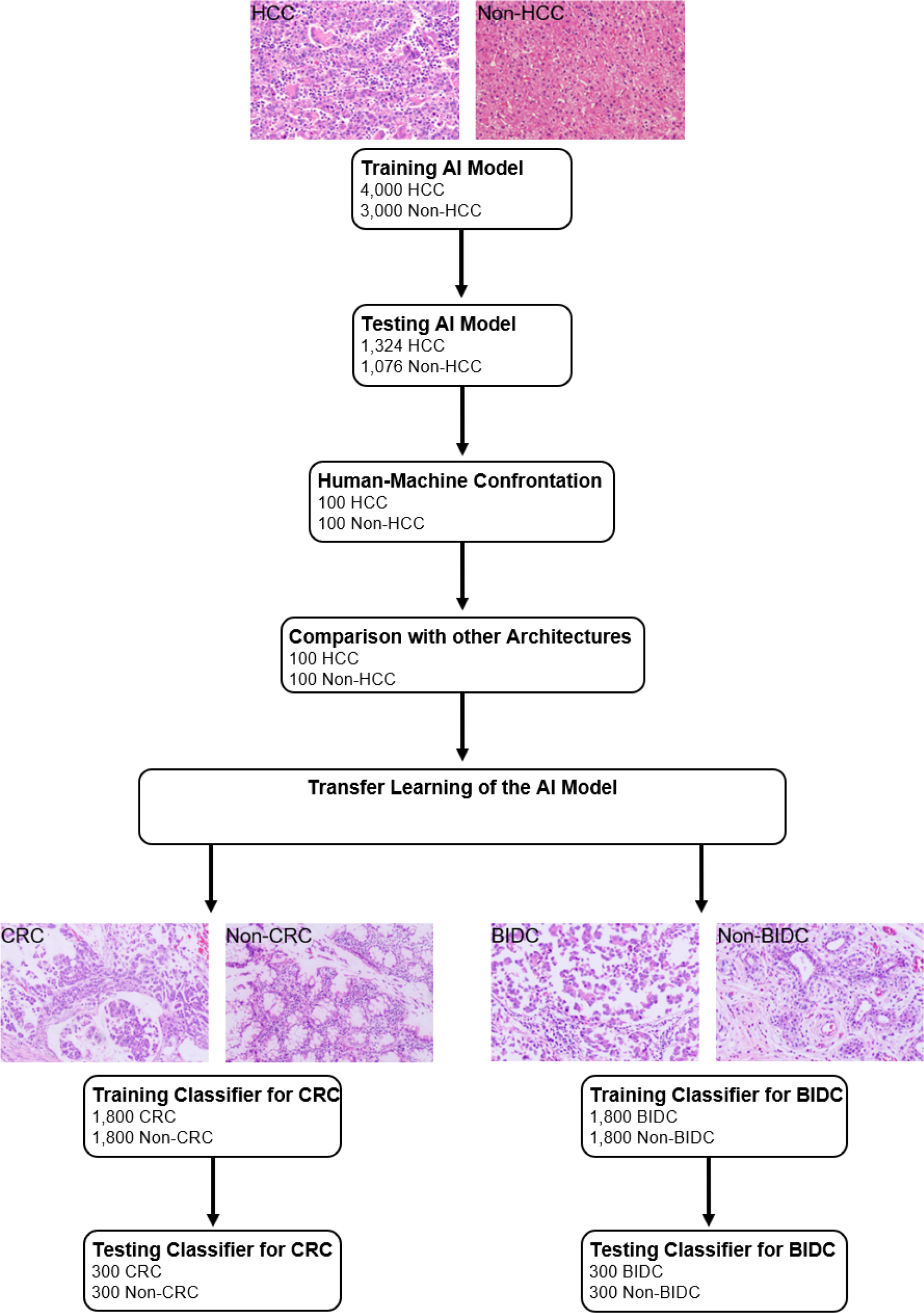
Study Design

### Statistical analysis

To evaluate the classification performance of the AI algorithm on histological images, three indies including accuracy, sensitivity, and specificity were calculated. The receiver operating characteristics (ROC) curves plot the true positive rate (sensitivity) versus the false positive rate (1-specificity). P < 0.05 was set as the level for statistical significance for two-tailed paired test.

## Results

### Patient and image characteristics

We obtained 7000 liver pathology slide images generated from 2745 patients enrolled from Xiamen Hospital of Traditional Chinese Medicine, where 4000 images showed confirmed HCC while the other 3000 images confirmed other diseases, such as CHB/CHC with or without cirrhosis. All these images passed an initial image-quality check, and were randomly divided into training set and validation set at a ratio of 3:1 to train and validate the classifying performance of the AI algorithm. This process was repeated five times.

### AI algorithm performance during process of training and validation

During training and validation process, accuracy and cross-entropy were plotted against the iteration step, which were shown in Supplementary Figure 2. By using the validation set as monitor, the mean sensitivity, specificity, and accuracy of the AI algorithm were calculated as 98.6%, 98.5%, and 98.5%, respectively.

### AI algorithm performance evaluated by test set

We also obtained 2400 images showing or not showing HCC, generated from 873 patients enrolled from Zhongshan Hospital Xiamen University, which were used to further evaluate the performance of the AI algorithm. In these 2400 images, 1324 of them showed HCC while 1076 of them showed non-HCC. By using the test set as monitor, the sensitivity, specificity, and area under ROC curve of the AI algorithm were calculated as 99.1%, 98.0%, and 0.960, respectively.

### Comparison between the results of the AI algorithm and human experts

To compare the performances of the AI algorithm and human experts, we chose another randomly selected set of 200 images consisting of 100 images with HCC and 100 images without HCC. All 200 images were sent to both the AI algorithm and human experts to make clinical decisions. The accuracy, sensitivity, and specificity for the AI algorithm were 98.5%, 98.0%, and 99.0%. For human experts, sensitivity ranged from 86.0% to 97.0% while specificity ranged from 91.0% to 100%.

Compared with human experts, the AI algorithm tended toward a more balanced performance between sensitivity and specificity. However, a remarkable variation was observed between sensitivity and specificity distinguishing it from the results by the human experts. The performances of the AI algorithm and human experts were presented in Figure 2.

**Figure 2.**
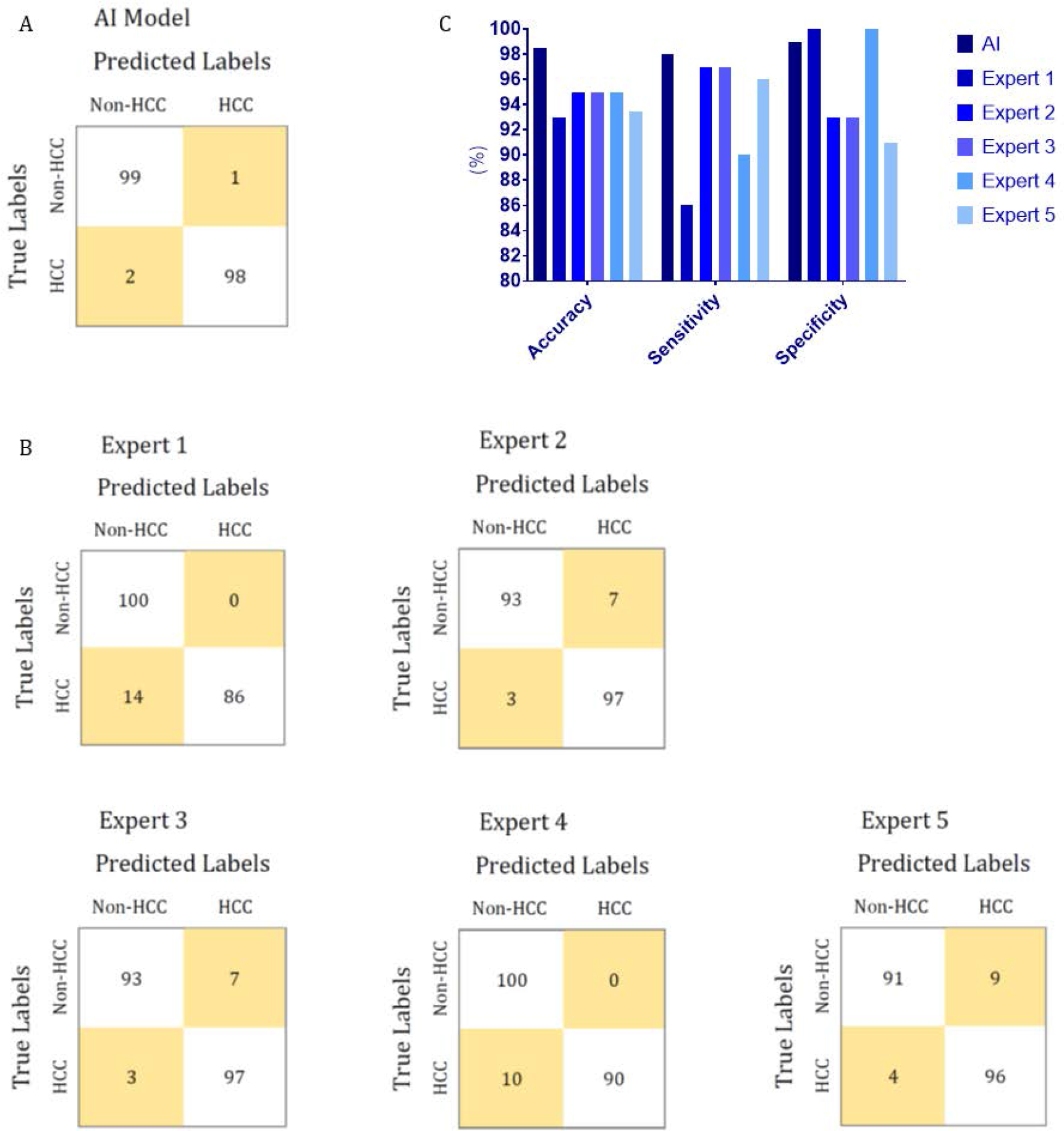
Performances of the proposed AI model and human experts during the human–machine comparison. A, Confusion matrix of the proposed AI model for HCC diagnosis; B, Confusion matrix of human experts for HCC diagnosis; C, Comparison of between the performance of the proposed AI model and that of human experts for HCC diagnosis.

### Comparison between the AI algorithm and other architectures

Two hundred images used for the human–machine comparison was also employed to compare the performance of the AI algorithm with those of the other architectures. The HCC image-recognition sensitivity, specificity, and accuracy of the proposed AI system was superior to those of AlexNet and GoogLeNet, as reported in Supplementary Table 1 and Supplementary Figure 3.

### Transfer learning of the AI algorithm to CRC

To evaluate the proposed transfer-learning performance of the AI system, 3600 colorectal-tissue microscope slide images from Xiamen Hospital of Traditional Chinese Medicine were collected to train and validate the AI algorithm. These 3600 images consisted of 1800 images showing CRC and another 1800 images not showing CRC. Another 600 colorectal-tissue microscope images from Zhongshan Hospital Xiamen University were also collected as a test set. As was shown in Figure 3, after limited training, the proposed AI algorithm showed excellent accuracy in CRC and non-CRC image classification based on transfer learning. An accuracy of 96.8% was achieved, with a sensitivity of 97.0% and a specificity of 96.7%.

**Figure 3.**
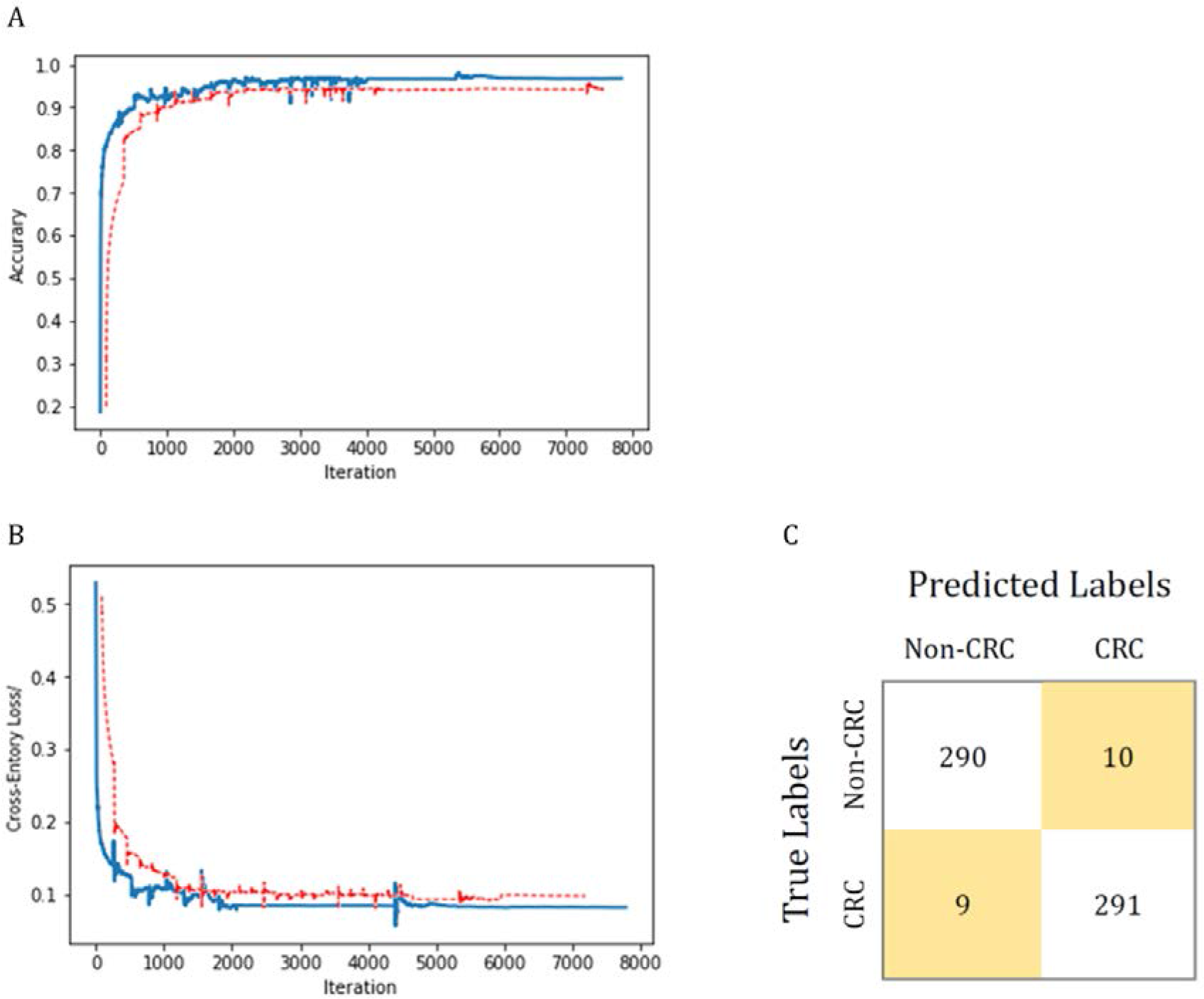
Transfer-learning performance of CRC diagnosis using colorectal tissue microscope slide images. In panel (A) and (B), the training dataset is shown in blue and the test dataset is shown in red. Accuracy is plotted against the iteration step (A), and cross-entropy loss is plotted against the iteration step (B) during the length of the training of the binary-class classifier over the course of 8000 steps. The curve is smoothed. The test accuracy and loss show better performance. Panel (C) shows the confusion matrix of best test image model classification. The model successfully classifies CRC separately from non-CRC.

### Transfer learning of the AI algorithm to BIDC

Microscope slide images from breast tissue were collected to further evaluate the transfer-learning performance of the proposed AI system. A total of 3600 histologic images of breast obtained from Xiamen Hospital of Traditional Chinese Medicine were employed to train and validate the AI algorithm. In these 3600 images, 1800 images showed BIDC while another 1800 images showed non-BIDC. Another 600 breast microscope images from Zhongshan Hospital Xiamen University were also obtained as a test set. As was shown in Figure 4, after training, the proposed AI system showed an accuracy of 96.0%, with a sensitivity of 95.7% and a specificity of 96.3%, in classifying images into BIDC or non-BIDC.

**Figure 4.**
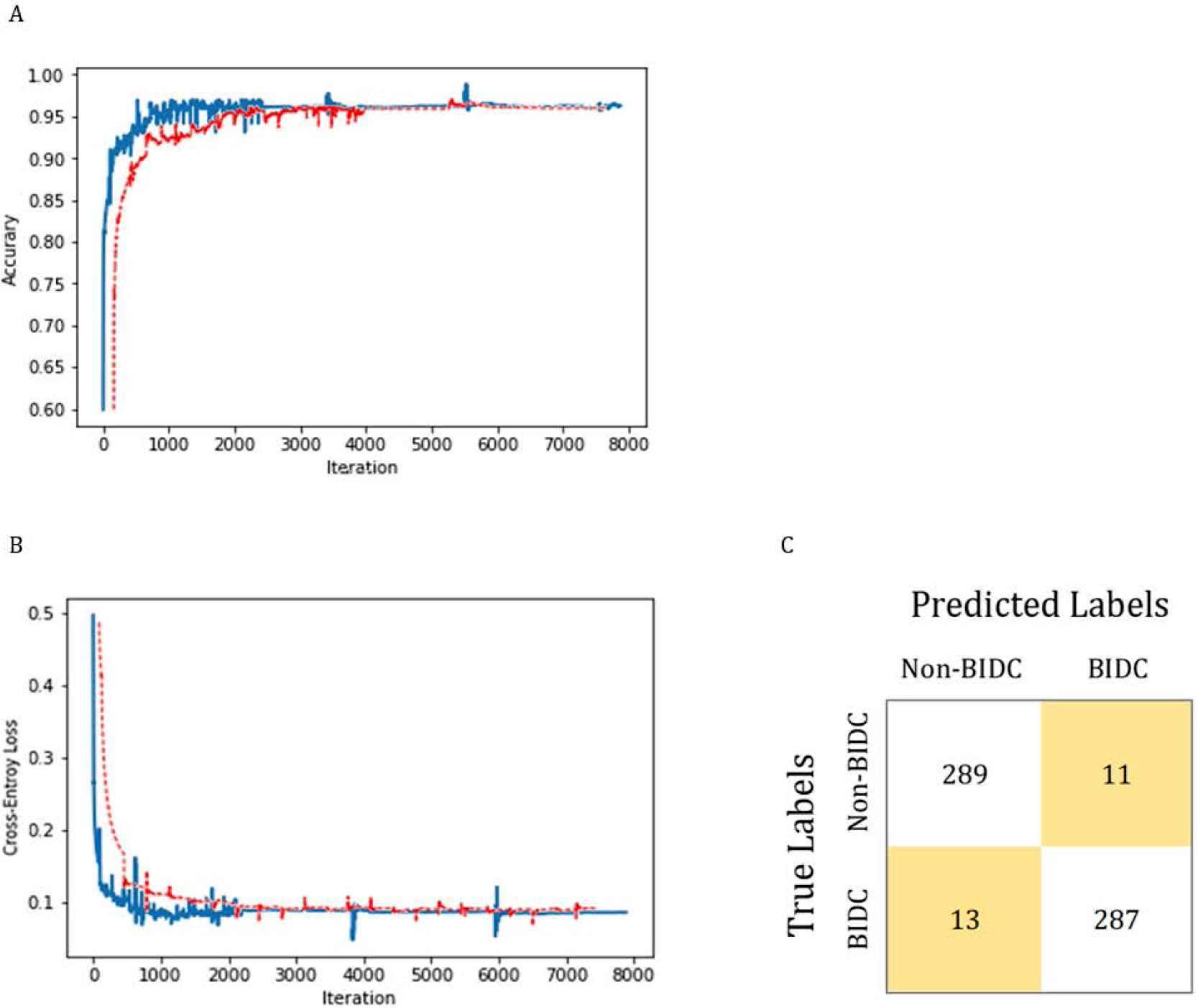
BIDC diagnosis transfer-learning performance using breast tissue microscope slide images. In panel (A) and (B), the training and test datasets are shown in blue and red, respectively. Classification accuracy is plotted against training epochs, and in (B), the categorical cross-entropy loss is shown as a function of training epochs for the binary classification problem. The curve is smoothed. Panel (C) shows the model-classification confusion matrix for test image classification. As shown, the proposed model successfully classifies BIDC from non-BIDC images.

## Discussion

In this paper, we described a general AI algorithm for the interpretation of histological images from the liver, colorectum, and breast. Although medical imaging, such as CT and MRI, are widely used for HCC diagnosis, CT and MRI detection is poor for HCCs < 1.0 cm, especially for the patients with cirrhosis.(22) For those patients without definitive findings on either CT or MRI, a biopsy may be a the only detection method.(1) Owing to potential interobserver bias that may be present when reviewing histological images generated from biopsy, AI may be a useful ancillary tool for HCC identification. In addition, the proposed model demonstrated competitive performance when analyzing liver histological images. This was accomplished without the need for a highly specialized, deep learning machine and without a very large training database. When the model was trained with 7000 images (3500 images for each class), high performance accuracy, sensitivity, and specificity were achieved for the correct diagnosis. Moreover, the model’s performance in diagnosing HCC was comparable to the diagnosis by experts with significant clinical experience in liver pathology.

By employing another 200 images (100 images for each class) as test set, the proposed AI model showed a more balanced performance between sensitivity and specificity in recognizing HCC compared with that of human experts. Accuracy of the proposed AI model was superior to that of experts, which also showed a remarkable variation between sensitivity and specificity. The test set as aforementioned was used for comparison between the proposed ResNet-34-architecture-based AI model and other AI architectures including AlexNet and GoogLeNet. The proposed AI model achieved superior accuracy, sensitivity, and specificity, thus demonstrating the robustness of the model.

In the study by Kermany et al, over 100000 OCT images were employed to train the AI framework.(14) However, only 3600 images were used to train our AI system in our study based on transfer learning, yet excellent diagnostic performance was achieved. Thus, even with a limited training dataset, the transfer-learning system demonstrated highly effective classifications. Transfer-learning techniques for image analysis could potentially be employed for a wide range of medical images across multiple disciplines. In fact, a direct illustration of its wide applicability in the analysis of two similar histological image types (CRC and BIDC) was shown. After a much smaller amount of training, the proposed AI model reported an accuracy of 96.8% and 96.0% for CRC and BIDC, respectively. Importantly, the proposed AI model showed a balance between sensitivity and specificity. Thus, the proposed AI system has potential universality in the classification of histological images.

An AI model trained from an extremely large training dataset would have superior performance to that of a transfer-learning-based model trained from a relatively small training dataset. However, in practice, *de novo* training of a convolutional neural network usually needs an unlimited supply of training data and would require weeks to achieve a good accuracy. By using the retraining layers from other medical classifications, a transfer-learning-based model would likely yield a highly accurate model in much less time. Thus, for difficult-to-collect medical images, transfer-learning-based image recognition is more practical. Given that imaging-based diagnosis has played a crucial role in guiding treatment, extending the proposed AI’s application to diagnoses and treatment recommendations is a promising area for future investigation.

## Data Availability

None

## List of Abbreviations

HCC: Hepatocellular carcinoma
CRC: colorectal cancer
BIDC: breast invasive ductal carcinoma
AI: artificial intelligence

## CONFLICTS OF INTEREST

### Guarantor of the article

Jin-Shui Pan, MD, PhD.

### Specific author contributions

Conception or design of the work: J.S.P., M.Z.H. and Q.Z.Z. Acquisition and analysis of data: Q.Z.Z., W.M.C., C.J.Z., Q.Q.X., F.Z., M.J.L., X.D., Q.Z.Z., and M.Z.H. Interpretation of data: M.F and J.S.P. Drafting of the manuscript: J.S.P. Revision for critically important intellectual content and final approval of the version to be published: J.S.P., M.Z.H. and Q.Z.Z.

### Financial support

This work was supported by the National Natural Science Foundation of China No.81871645 (J.S.P.). The funding sources did not have any role in the design and conduct of the study; collection, management, analysis, and interpretation of the data; preparation, review, or approval of the manuscript; and decision to submit the manuscript for publication.

## Acknowledgements

We are very grateful to the enrolled patients who provides precious histological images.

## Supplementary Tables

**Supplementary Table 1.** HCC image-recognition performance of the AI model and other architectures

|  | Sensitivity (%) | Specificity (%) | Accuracy (%) |
| --- | --- | --- | --- |
| AlexNet | 97.0 | 95.0 | 96.0 |
| GoogLeNet | 98.0 | 96.0 | 97.0 |
| Our AI Model | 99.0 | 98.0 | 98.5 |

**Supplementary Table 2.**
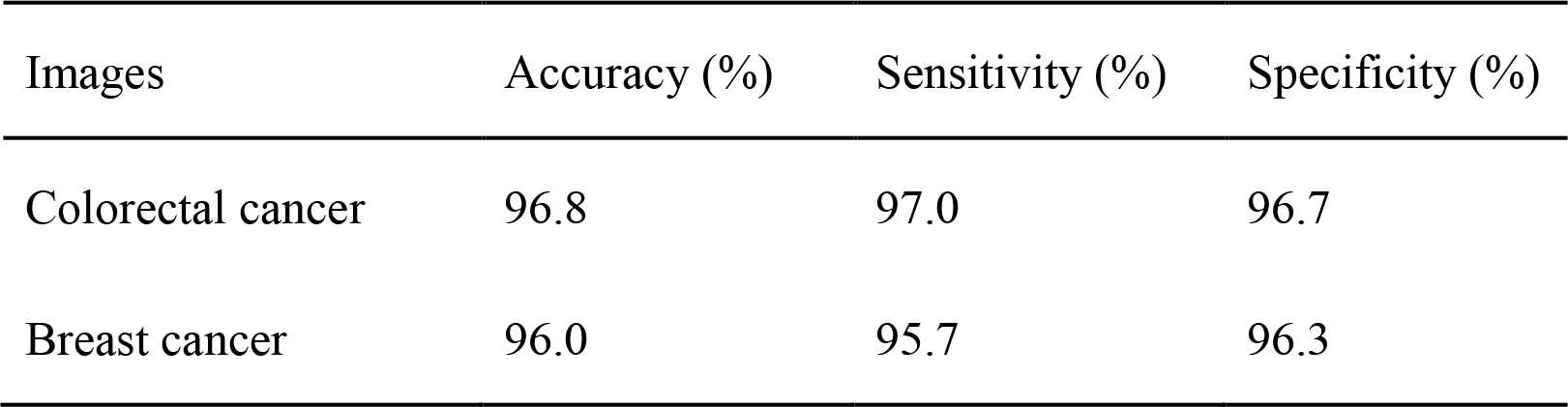
Proposed AI framework performance in classifying other types of medical images by using transfer learning

| Images | Accuracy (%) | Sensitivity (%) | Specificity (%) |
| --- | --- | --- | --- |
| Colorectal cancer | 96.8 | 97.0 | 96.7 |
| Breast cancer | 96.0 | 95.7 | 96.3 |

## Supplementary Figures

**Supplementary Figure 1:**
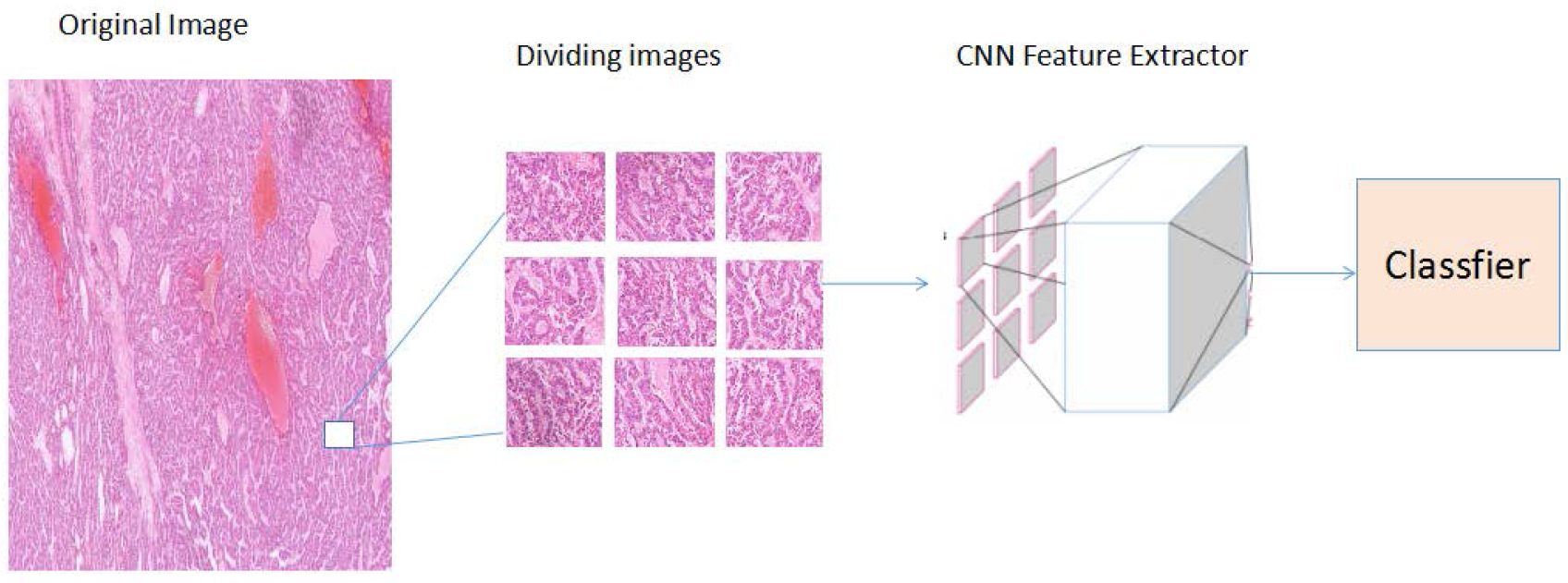
Algorithm Architecture Whole slide images were divided into n images, which were sent to the CNN feature extractor. Finally, the image features were determined by the constructed classifier.

**Supplementary Figure 2:**
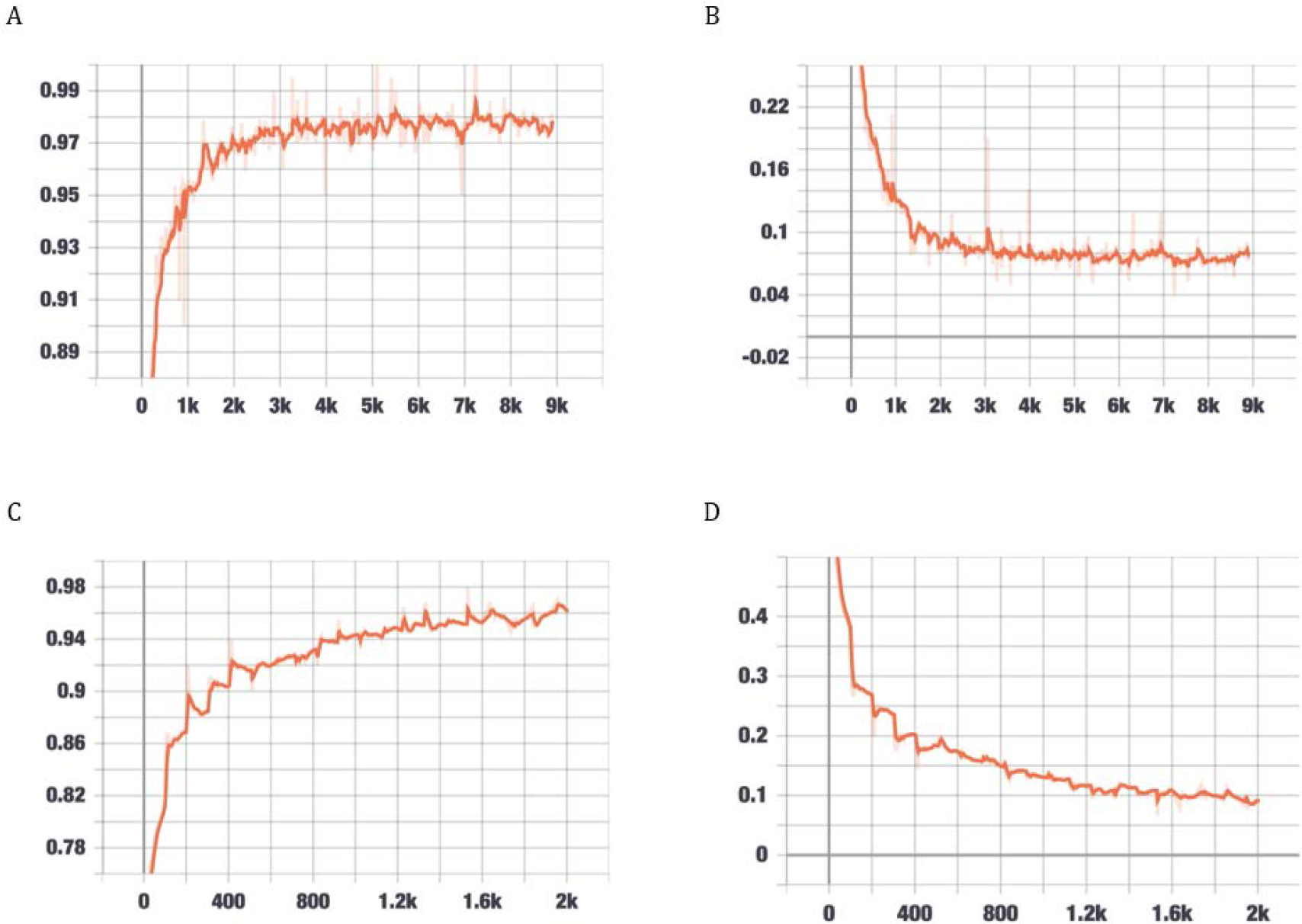
Performance of HCC recognition during validation. (A) Classification accuracy is plotted against training epochs, and in (B), the categorical cross-entropy loss is shown as a function of training epochs for the binary classification problem. (C) Classification accuracy is plotted against validation epochs, and in (D), the categorical cross-entropy loss is shown as a function of validation epochs for the binary classification problem. The curve is smoothed.

**Supplementary Figure 3.**
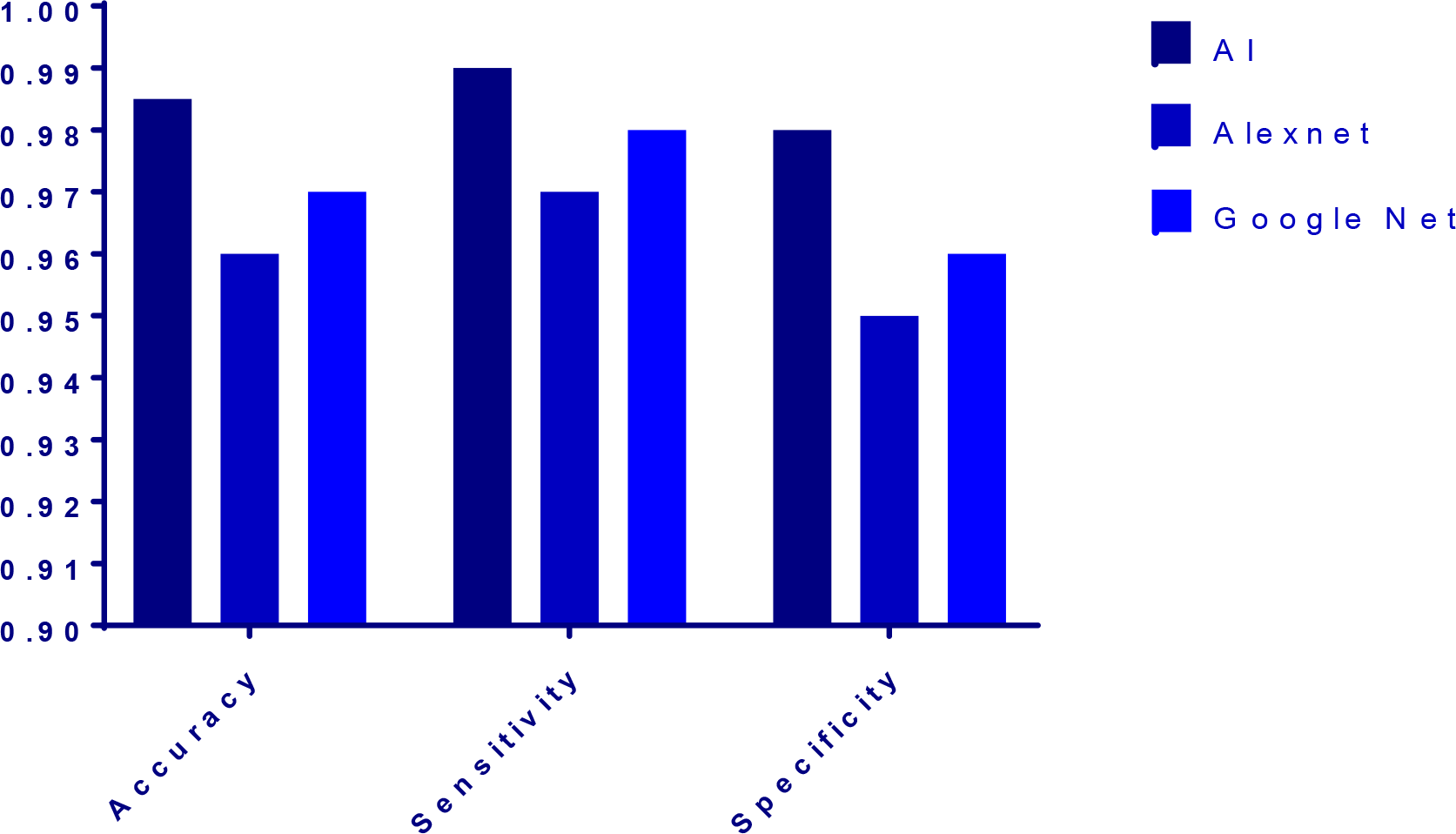
Performances of the proposed AI model and other architectures for HCC diagnosis.

